# A three gene signature predicts response to selinexor in multiple myeloma

**DOI:** 10.1101/2022.02.25.22271401

**Authors:** Paula Restrepo, Sherry Bhalla, Adolfo Aleman, Violetta Leshchenko, David T Melnekoff, Sarita Agte, Joy Jiang, Deepu Madduri, Joshua Richter, Shambavi Richard, Ajai Chari, Hearn Jay Cho, Sundar Jagannath, Christopher J Walker, Yosef Landesman, Alessandro Laganà, Samir Parekh

**Affiliations:** Tisch Cancer Institute, Icahn School of Medicine at Mount Sinai, New York, NY, United States; Department of Oncological Sciences, Icahn School of Medicine at Mount Sinai, New York, NY, United States; Department of Hematology & Medical Oncology, Icahn School of Medicine at Mount Sinai, New York, NY, United States; Department of Genetics & Genomic Sciences, Icahn School of Medicine at Mount Sinai, New York, NY, United States; Graduate School of Biomedical Sciences, Icahn School of Medicine at Mount Sinai, New York, NY, United States; Janssen Pharmaceutical Research & Development, Raritan, NJ, United States; Multiple Myeloma Research Foundation, Norwalk, CT, United States; Karyopharm Therapeutics, Newton, MA, United States

**Author notes:** **Address for Correspondence:** Dr. Alessandro Laganà, Department of Genetics and Genomic Sciences, Icahn School of Medicine at Mount Sinai, New York, NY, USA, Dr. Samir Parekh, Division of Hematology and Medical Oncology, Tisch Cancer Institute, Icahn School of Medicine at Mount Sinai, New York, NY, USA.

**Keywords:** Drug sensitivity, Gene expression profile, multiple myeloma, Survival prediction

## Abstract

Selinexor is the first selective inhibitor of nuclear export (SINE) to be approved for treatment of relapsed or refractory multiple myeloma (MM). There are currently no known genomic biomarkers or assays to help select MM patients at higher likelihood of response to selinexor. Here, we aim to characterize transcriptomic correlates of response to selinexor-based therapy, and present a novel, three-gene expression signature that predicts selinexor response in MM. We analyzed RNA sequencing of CD138^+^ tumor cells from bone marrow of 100 MM patients who participated in the BOSTON study and identified three genes upregulated in responders. Then, we validated this gene signature in 64 patients from the STORM cohort of triple-class refractory MM, and additionally in an external cohort of 35 patients treated in a real world setting outside of clinical trials. We also found that the signature tracked with response in a cohort of 57 patients with recurrent glioblastoma treated with selinexor. Furthermore, the genes involved in the signature, *WNT10A, DUSP1*, and *ETV7*, reveal a potential mechanism through upregulated interferon-mediated apoptotic signaling that may prime tumors to respond to selinexor-based therapy. This signature has important clinical relevance as it could identify cancer patients that are most likely to benefit from treatment with selinexor-based therapy.

## INTRODUCTION

Selinexor is the first selective inhibitor of nuclear export (SINE) approved for treatment of relapsed or refractory multiple myeloma (MM) and diffuse large b-cell lymphoma (DLBCL).^1, 2^ This approval is well supported by recent clinical trial data, most notably the STORM and BOSTON studies. In the STORM phase II clinical trial, oral selinexor and dexamethasone were administered twice weekly in patients with triple-class refractory MM, with 26% of patients exhibiting a partial response (PR) or better, and a median progression-free survival (PFS) at 3.7 months.^1, 3^ The phase III BOSTON trial compared the efficacy of once weekly selinexor in combination with once weekly bortezomib and dexamethasone with standard twice weekly bortezomib and dexamethasone in patients with previously treated MM and found an overall response rate (ORR) of 76.4%, and a median PFS of 13.9 months for patients receiving the novel treatment regimen, in comparison to 9.5 months for those receiving the standard treatment.^1^

Mechanistically, selinexor binds to the karyopherin exportin 1 (XPO1), which is responsible for shuttling more than 200 oncogenic and tumor suppressor proteins and mRNA transcripts to the cytoplasm.^4^ This inhibition of nuclear export, the sequestration of tumor suppressor proteins in the nucleus, and the prevention of select oncogene mRNA translation into oncoproteins ultimately induces cancer cell death while permitting the survival of non-malignant cells.^5–7^ Selinexor has a unique adverse event profile due to its novel mechanism of action, and adverse events (AE) are common, with severe AE reported in 52% of patients in the BOSTON trial, and 80% of patients in the STORM trial.^1, 3^ Identifying biomarkers that help predict treatment responses and toxicity is essential to targeted selinexor-based therapeutic intervention.

Systematic approaches to biomarker discovery for selinexor response that leverage next-generation sequencing are generally lacking in the literature. While MM cells tend to over-express XPO1 compared to normal plasma cells, XPO1 alterations have not correlated significantly with response to treatment with SINE compounds with the exception of a few *in-vitro* studies.^4–6, 8^ The STORM study presented a brief transcriptomic analysis that identified a potential four-gene signature based on imputed protein activity.^3^ A few candidate biomarkers identified through bioinformatics analyses have also been reported in conference proceedings.^9, 10^ However, limitations of these studies include small sample sizes and lack rigorous validation with external cohorts of selinexor-treated patients.

Here, we analyzed RNA sequencing data from 256 patients from multiple studies of selinexor-based therapy to characterize transcriptomic correlates of response to selinexor. We discovered a novel three-gene signature predicting response to selinexor using RNA-seq data from the BOSTON study and validated it on data from the STORM clinical trial and on an additional independent cohort of patients treated with selinexor at the Mount Sinai Hospital in “real-world” conditions outside of a clinical trial. The three-gene signature is biologically interpretable and opens a path for evaluating a mechanism of response in future studies. More importantly, this signature has the potential to identify patients most likely to benefit from treatment with selinexor-based therapy, ultimately reducing toxicities and improving outcomes.

## RESULTS

### Patient characteristics and transcriptomic profiling of selinexor-treated MM tumors

#### BOSTON

We performed RNA-seq on CD138^+^ bone marrow plasma cells from 100 patients who participated in the BOSTON study (Table 1). We included 53 samples from the selinexor, bortezomib, and dexamethasone (XVd) arm of the study, and 47 samples from the bortezomib and dexamethasone (Vd) arm. In the XVd arm, most patients were male (60.3%), compared with 46.8% in the Vd arm. In both study arms, and across the entire cohort, the median age was 67 years. The ORR in the XVd arm, defined as partial response (PR) or better, was 75.47%, compared to an ORR of 65.95% in the Vd Arm (Table 1). These observations suggest that the addition of selinexor to a regimen of bortezomib and dexamethasone in MM offers a clinical benefit over bortezomib and dexamethasone alone, and are consistent with the findings of previous studies, including the main BOSTON trial.^1^

**Table 1.**
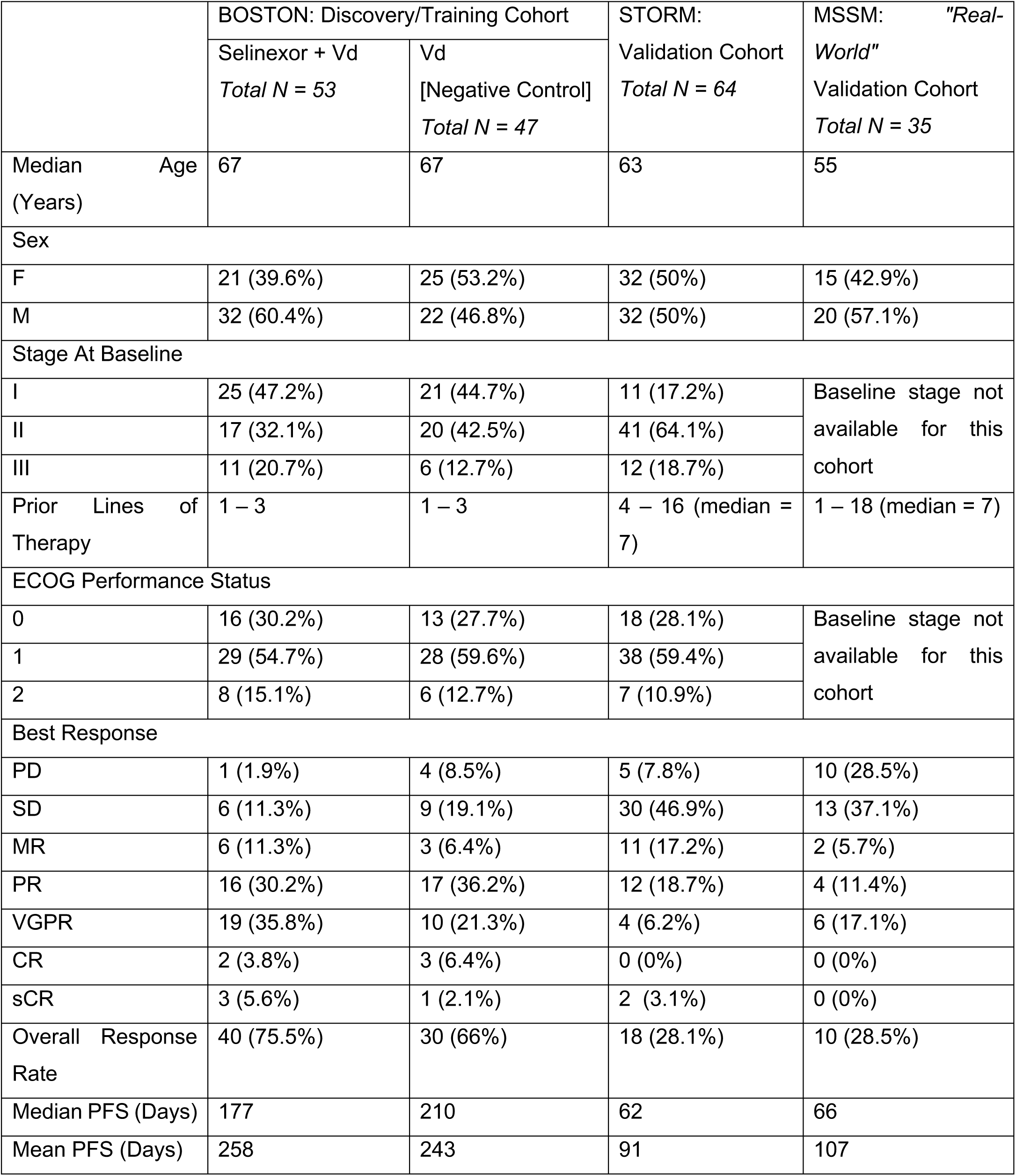
Cohort Clinical Characteristics.

For validation purposes, RNA-sequencing on CD138^+^ selected cells from two external cohorts were leveraged: MM patients treated with selinexor-dexamethasone who participated in the STORM trial (N = 64), and MM patients treated post-FDA approval at Mount SInai Hospital outside of any clinical trial setting (MSSM cohort, N = 35).^3^ Patients in the STORM cohort had failed at least five prior lines of therapy.^3^ The median PFS was 62 days, and 18 patients (28.1%) achieved a response of PR or better (Table 1). Patients in the MSSM cohort received a median of 7 lines of therapy (range = 1-18) prior to starting selinexor based therapy, which was often administered in combination with a variety of other agents. The median PFS was 66 days, and 10 patients (28.5%) achieved a response of PR or better (Table 1). The most common combination drug strategy in addition to the selinexor backbone was carfilzomib and dexamethasone, used in 7 of 35 (20%) patients.

We performed principal component analysis (PCA) on the normalized, batch-corrected expression matrix to understand sources of variation in the BOSTON, STORM, and MSSM datasets, which did not identify any specific bias (Fig S1). Overall, we did not find any significant or precise predictors of response to selinexor from clinical, demographic, or cytogenetic markers.

### Differential expression analysis identifies genes associated with selinexor response

The strategy used for differential expression (DE) analysis is summarized in Fig 1A. To identify genes whose expression is associated with longer PFS or better depth of response to selinexor, we performed a series of DE comparisons across 9 unique different PFS or depth of response (defined according to International Myeloma Working Group [IMWG] criteria) thresholds in both the XVd and Vd arms (Fig S2, Table S1).^11^ Across all response thresholds, there were a total of 107 unique significant DE genes between better and worse responders in the XVd Arm (27 upregulated, 70 downregulated, FDR < 0.05) and 560 unique DE genes in the Vd Arm (398 upregulated, 162 downregulated, FDR < 0.05). To identify genes whose association with PFS- or IMWG response category in the XVd arm was not also associated with bortezomib/dexamethasone therapy, we retained for further analysis genes that were significant in the XVd arm but not in the Vd arm for each corresponding cutoff. In total, there was a moderate overlap (up to 31%) across genes identified through the different comparisons, with 6 out of 24 (25.0%) uniquely downregulated genes and 12 of 33 (36.26%) uniquely upregulated genes overlapping across at least two different cutoffs.

**Fig 1.**
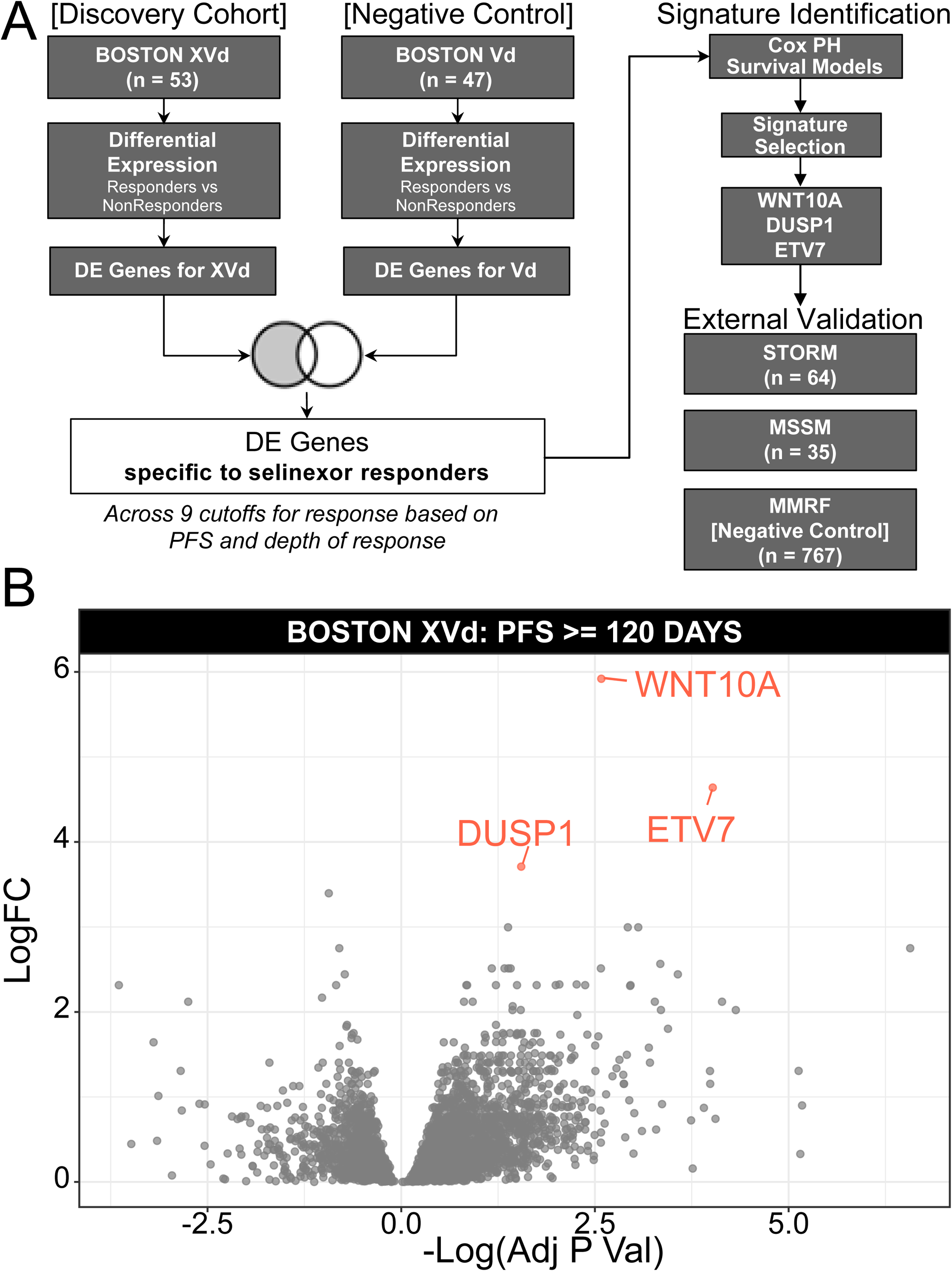
Differentially expressed genes in selinexor responders. A) Experimental design for identifying DE genes specific to selinexor. B) Volcano plot of the BOSTON XVd DE comparison with a PFS ≥ 120 days response cutoff showing the three significantly upregulated genes used to generate the three-gene response signature.

### A three-gene signature predicts selinexor response

Next, we identified a gene expression signature for selinexor response. Each DE gene set identified through the 9 unique comparisons was split into upregulated and downregulated subsets based on logFC in the XVd Arm, resulting in 17 candidate gene signatures for further analysis (9 upregulated, 8 downregulated). We performed Gene Set Variation Analysis (GSVA) on the normalized and batch-corrected expression matrix to calculate a unique score for each of the 17 candidate gene signatures. For each signature, a univariate proportional hazard model was generated in the XVd arm. The gene signature with the best performance in model training was identified based on a ranking procedure using repeated four-fold cross validation and the spearman correlation of the signature with PFS (Fig 2A; see Methods).

**Fig 2.**
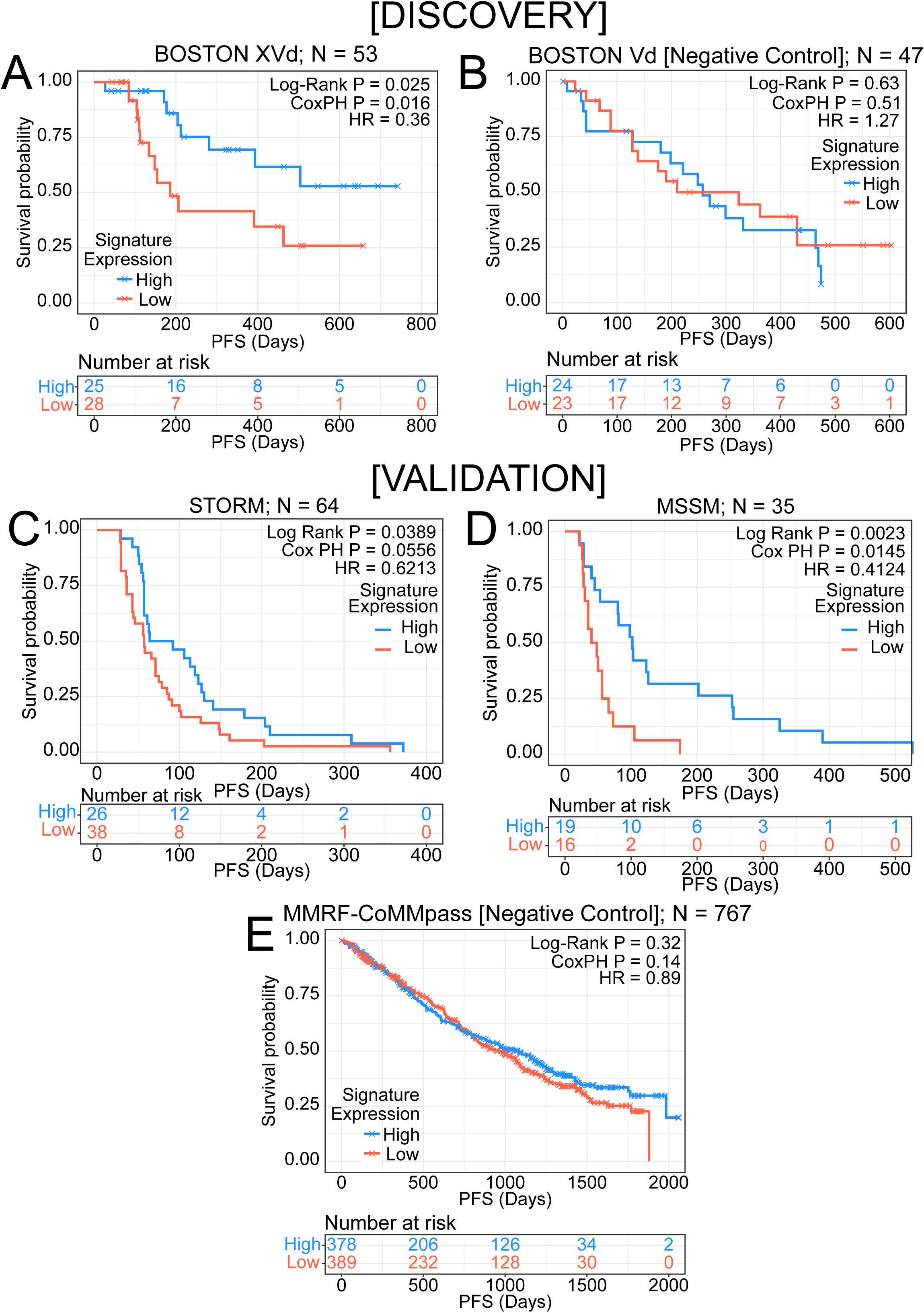
Upregulation of the signature is associated with longer PFS in pre-selinexor-treated MM. A) Kaplan-Meier curve of low versus high expression of the three gene GSVA signature in the XVd arm of the BOSTON cohort shows a significant association with longer PFS in patients with upregulation of the signature. B) Using the Vd arm as a negative control, the three gene signature is not associated with PFS. C) Three gene signature validates in the STORM cohort of triple-class refractory MM patients who received selinexor as a single-agent treatment. D) The signature also validates in 35 patients that received selinexor in a retrospective cohort at MSSM that is not part of a clinical trial and is a more heterogeneous patient population reflective of “real world” treatment settings. E) Signature does not validate in non-selinexor, standard of care treated MMRF-COMMPASS samples.

The best performing signature was composed of three genes, *WNT10A, DUSP1*, and *ETV7*. It was correlated with PFS (Spearman rho = 0.46, P = 0.0007) and was upregulated in XVd patients with PFS ≥ 120 days. The signature was predictive when using a proportional hazard model (FDR=0.047, HR=0.36 [95% CI = 0.14-0.84]; log-rank P = 0.017, Fig 2A). To further ensure that the predictive effect of the signature was not due to random variations, we also performed a permutation test and found that the signature was more predictive than a GSVA score composed of three randomly selected genes (10,000 permutations, P = 0.03; see methods). In patients who achieved a partial response (PR) or better, higher signature expression was significantly associated with longer duration of response (DOR), defined as the number of days from IMWG partial response to progression (Fig S2A). Finally, an ordinal regression found that higher signature expression was significantly associated with deeper IMWG response category (P = 0.0247, R^2^ = 0.095, Spearman Rho = 0.317, Fig 3A), suggesting that the signature is associated with both duration and depth of response.

**Fig 3.**
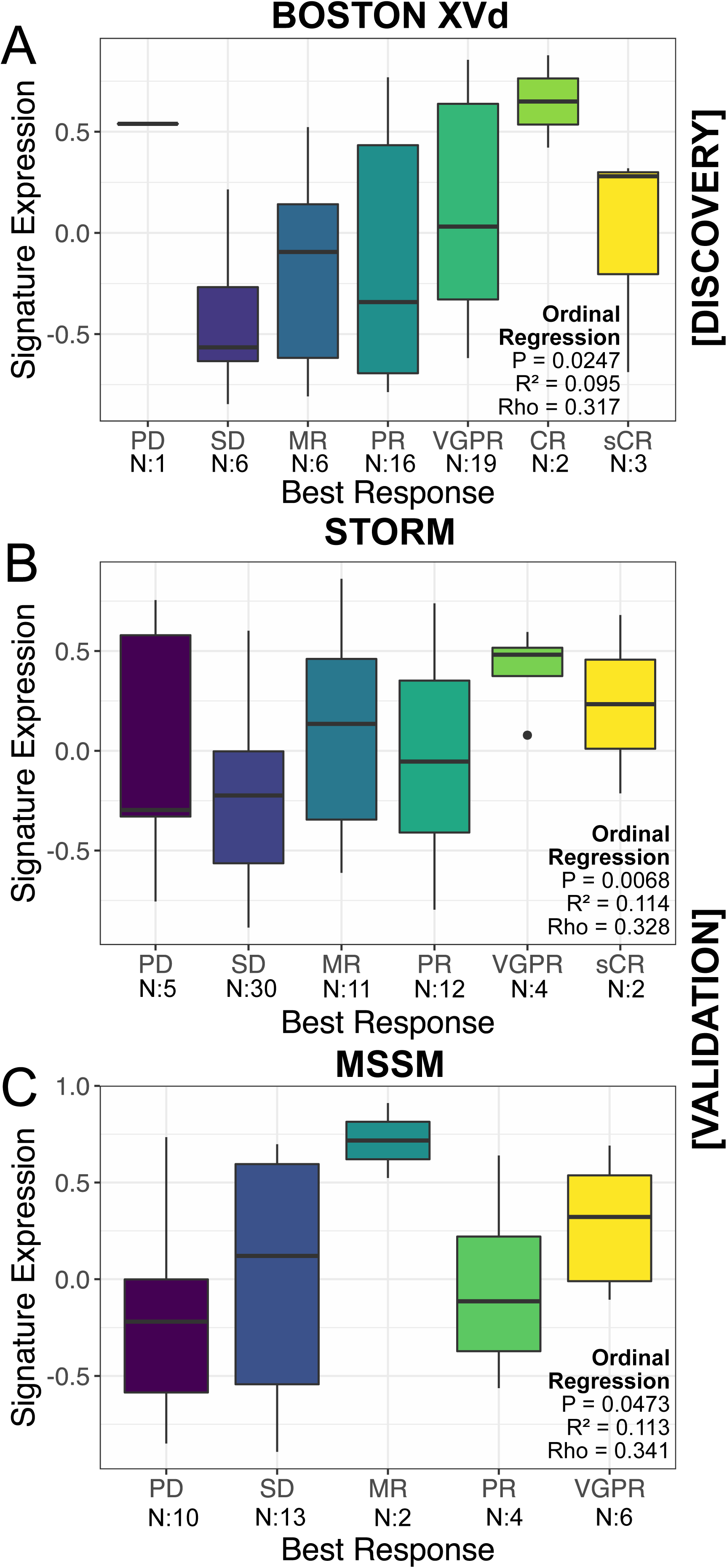
Higher signature expression is significantly associated with better depth of response. Higher three-gene signature expression is significantly associated with better IMWG response category via ordinal regression modeling for patients in the A) BOSTON XVd, B) STORM, and C) MSSM cohorts.

The same analysis was applied to the Vd arm as a negative control under the rationale that a signature specific to selinexor response would not accurately distinguish long or short PFS in cohorts treated with non-selinexor based therapy. The three-gene signature did not track with PFS or DOR in the Vd arm, as shown in Fig 2B and Fig S2B.

### The three-gene signature is validated in external cohorts

#### STORM

A log-rank test comparing signature expression higher or lower than the cutoff of zero in the STORM dataset also validated the finding from BOSTON that higher signature expression is associated with PFS (log rank P = 0.039; N=64). We found that the linear association with PFS and the signature performed nominally well, despite not reaching statistical significance (N=64; P = 0.056, HR=0.621 [95% CI = 0.483-0.389], Fig 2C). The poorer signature performance in a cox regression may be explained by lower PFS, as patients in the STORM cohort were triple-class refractory with more advanced disease to begin with. An ordinal regression testing the association between signature expression and depth of response also validated in the STORM cohort (P = 0.0068, R^2^ = 0.114, Spearman Rho = 0.328, Fig 3B).

#### MSSM

Using cox regression, we found that higher expression of the three-gene signature was significantly associated with survival (P = 0.0145, HR = 0.41 [95% CI = -0.467-0.551]). This result was also replicated via log rank testing (P = 0.0023, Fig 2D). Furthermore, we found a statistically significant correlation of the gene expression signature with PFS via Spearman correlation (rho = 0.4, P = 0.01). Higher expression of the signature was also significantly towards a better IMWG response category via ordinal regression (P = 0.0473, R^2^ = 0.113, Spearman Rho = 0.341, Fig 3C).

#### MMRF-CoMMpass

We additionally used the MMRF-CoMMpass dataset (N=767) as a negative control and found that the signature was not predictive of PFS in patients who were treated with non-selinexor based, standard of care therapies. The signature did not show a significant association with progression free survival via cox regression (P = 0.14, HR = 0.89 [95% CI = 0.73-1.04]) or via log rank test (P = 0.32, Fig 2E). Taken together, these results suggest that the signature is specific to selinexor treatment response and is not reflective of overall prognosis.

#### KING Study in Recurrent Glioblastoma

Lastly, we sought to test whether the signature would be predictive in cohorts of patients with other cancer types treated with selinexor from other cancer types. To test this hypothesis, we obtained RNA-seq of tumor samples from 57 patients with recurrent glioblastoma who were treated with selinexor monotherapy as part of the phase II KING trial (NCT01986348).^12^ Overall, we found that patients with higher expression of the signature experienced improved PFS, although statistical significance was not achieved (log rank P = 0.078, Cox PH P = 0.0734, HR = 0.549, Fig S3A). However, patients who achieved a clinical benefit with a partial response (PR) or better had significantly higher signature expression (Wilcoxon P = 0.0034, Fig S3B).

### Gene-set enrichment analysis reveals response-associated activation in wnt, apoptosis, and mapk signaling pathways

We next identified pathways associated with response to selinexor in the BOSTON XVd cohort using gene set enrichment analysis (GSEA) on the MSigDB Hallmark gene sets.^13^ Notably, we found downregulation of MYC targets, as well as upregulation in KRAS, apoptotic, and interferon signaling among significantly enriched pathways in XVd responders (Fig S4).

## METHODS

### Patient Selection & RNA Sequencing

#### BOSTON and STORM

CD138^+^ cells were purified from bone marrow aspirates obtained from 100 patients who participated in the BOSTON study and 64 patients who participated in the STORM study. RNA extraction was performed using Qiagen Allprep RNA mini kit and library preparation was performed with the TruSeq Stranded mRNA (non-FFPE compatible) kit. For samples where the RNA quality was low, the Smart-Seq V4 Ultra Low Input Nextera XT kit was used. Total RNA sequencing was performed with 100 bp reads using an Illumina HiSeq 2500 instrument.

#### MSSM

The 35 patients were physician-referred as part of the multiple myeloma banking protocol approved by the Mount Sinai institutional review board (IRB). Informed written consent was obtained from each patient. CD138^+^ cells were isolated from bone marrow aspirates obtained prior to the start of selinexor-based therapy, and RNA isolation and sequencing was performed as previously described.^14^

#### MMRF-CoMMpass

Gene-level counts for 767 RNA-seq samples from the MMRF-CoMMpass dataset were downloaded from dbGaP (accession #phs00748).

#### KING

Gene-level read counts were obtained from pretreatment tumors from 57 patients with recurrent glioblastoma that were enrolled in the phase 2 KING study.^12^

#### Bioinformatics Processing

Raw reads were aligned to the GRCh38 human reference genome using STAR.^15^ Gene-level counts, obtained through featureCounts, were filtered to remove immunoglobulin and ribosomal transcripts, and to remove genes whose counts across all samples had zero variance.^16, 17^ Counts were converted to Log_2_CPM, normalized with voom, and corrected for batch effects or covariates identified through variancePartition analysis using the sva ComBat package.^17–19^

### Differential Expression

DE genes were identified in the BOSTON XVd and the Vd arm separately using DESeq2.^20^ Within each arm, DE genes relative to responders versus non-responders were generated across a total of 9 response cutoffs based on PFS, ORR, or a combination of both. For each response comparison, genes were designated as DE in selinexor responders if they were significantly DE in the XVd Arm (P_Adj_ < 0.05) and were not significantly DE in the corresponding comparison within the Vd arm (P > 0.05). DESeq2 analysis was performed on unfiltered raw counts per the tool’s requirements.^20^ Gene set enrichment analysis was applied to the selinexor-specific DESeq2 results in the XVd arm using the curated Hallmark and Reactome signatures publicly available from MSigDB using the fGSEA package.^13, 21^

### Survival Analysis

Each DE gene set identified through the 9 comparisons was split into upregulated and downregulated subsets based on logFC in the XVd Arm, resulting in 17 candidate gene signatures for further analysis (9 upregulated, 8 downregulated). Gene set variation analysis (GSVA) scores were calculated for all samples for the 17 candidate gene signatures from the covariate-normalized expression matrix.^22^ For each candidate gene signature, a univariate Cox proportional hazard model was generated in the BOSTON XVd arm. The gene signature with the best model performance in the discovery cohort was selected by ranking the highest Somer’s *D*_*xy*_ after repeated four-fold cross validation (n_*repeats*_ = 1000). The gene signature was also evaluated with a Cox proportional hazard model in the Vd arm as a negative control to ensure that performance was specific to selinexor. A permutation test was performed to evaluate whether the selected gene signature is more significant via log-rank Kaplan-Meier testing than a GSVA score composed of three randomly selected genes.

All validation tests were executed by first performing quantile-normalization of the expression matrix with the distribution of expression in the BOSTON dataset using feature-specific quantile normalization, followed by calculation of a GSVA score for the gene signature.^22, 23^ Survival was tested between groups that had low expression versus high expression, using a cut-off of zero, with a log-rank test and univariate cox regression carried out with the same procedures used for the discovery cohort.

## DISCUSSION

Selinexor is approved as a second-line therapy for MM and its efficacy is well supported by clinical trials. However, there are currently no known biomarkers to better guide selection of patients whose tumors are more sensitive to selinexor-based therapy. Furthermore, while the mechanism of action is well characterized for selinexor, little is known about the correlates of response or resistance to selinexor-based therapy. Here, we describe the transcriptomic characteristics of patients who respond to selinexor therapy. Further, we report the discovery of a robustly validated three-gene signature that is predictive of response to selinexor-based therapy in MM.

There is little literature to date on patient populations describing correlates to response in the context of selinexor therapy. Some studies have shown candidate biomarker activity in miRNAs as regulators of XPO1 and its targets, and certain mutations in *XPO1* and *XPO5* as prognostic markers for survival, but they have not been correlated to SINE drug sensitivity.^8, 24–27^ There are few studies that have explored biomarkers in selinexor therapy, and fewer with validation in external cohorts. One notable study found a signature based on imputed activity comprising four master regulator proteins, *IRF3, ARL2BP, ZBTB17*, and *ATRX*, but did not provide any external cohort validation data.^3^

We report here a gene expression signature composed of the combined upregulation of three genes, *ETV7, WNT10A*, and *DUSP1*, that precisely and accurately predicts both depth and duration of response to selinexor-based therapy in patients with MM. Furthermore, we present robust external validation and negative controls. This is the first robustly validated signature for selinexor response to date. Given the high rate of adverse events in selinexor-based therapy, the discovery of a predictive gene signature holds tremendous potential for biomarker-guided selection of candidates for selinexor-based therapy that are more likely to respond. Since the signature was validated in multiple heterogeneous patient cohorts, including selinexor-dexamethasone monotherapy (STORM), and in various combination drug scenarios outside of clinical trials (MSSM), it is both flexible and applicable to a wide variety of real-world scenarios where selinexor may be given in combination with other drugs. One limitation of the signature is that, since it relies on a GSVA score, its accuracy is dependent on a cohort with multiple samples and prediction improves with greater sample sizes. However, since it is composed of just three genes, it also holds potential for fast and simple implementation in clinical settings, potentially via qPCR-based assay development strategies or in combination with *ex-vivo* drug sensitivity assays. Furthermore, since the signature uses gene expression directly, it is more interpretable and easier to implement than previously cited signatures, which used imputed protein activity and are not as thoroughly validated.^3^

Interferon signaling is responsible for the anti-viral immune response and has anti-proliferative properties. It has been shown to play an important role in apoptotic signaling in MM, mostly through *IRF4, MYC*, and *BCL6*.^28–30^ Here, we found a strong enrichment for upregulated interferon signaling and genes involved in interferon signaling in patients who responded to XVd therapy. We also found a corresponding enrichment of dysregulated *MYC* and apoptotic signaling. Interferon signaling has been found to modulate response to XPO1 inhibition by eltanexor to treat viral infection.^31^ Additionally, *ETV7* has been identified as an interferon-stimulated gene.^32^ Based on these results and existing literature, we hypothesize that there may be a potential biological rationale for further evaluating the role of interferon signaling in MM selinexor response.

In conclusion, we report a novel gene expression signature for response to selinexor-based therapy in patients with MM. We have validated our findings in several external transcriptomic datasets of MM patients treated with selinexor-based regimens. Ongoing *in vitro* and mechanistic studies will help determine whether they are causative of response or simply a correlative readout of some other selinexor response mechanism. This signature has important clinical significance as it could identify patients most likely to benefit from treatment with selinexor-based therapy, especially in earlier lines of therapy.

## Data Availability

Raw sequencing data will be deposited in SRA upon publication. The code necessary to calculate selinexor signatures is provided in a GitHub repository at https://github.com/Lagana-Lab/selinescore.

## AUTHOR CONTRIBUTIONS

PR performed most of the downstream bioinformatics analysis, with assistance from SB and AL. PR, DTM, and DB processed the raw sequencing data. SA, AA, and VL, compiled clinical data for analysis. CW and YL provided data from the KING study. AC, SJ, SR, SP, DM, JR, HJC, YL provided patient samples. VL and AA performed library preparation for sequencing. PR wrote the initial manuscript with assistance from JJ. AL, SP, CW and YL conceptualized and supervised the study.

## FIGURES AND TABLES

**Fig S1.**
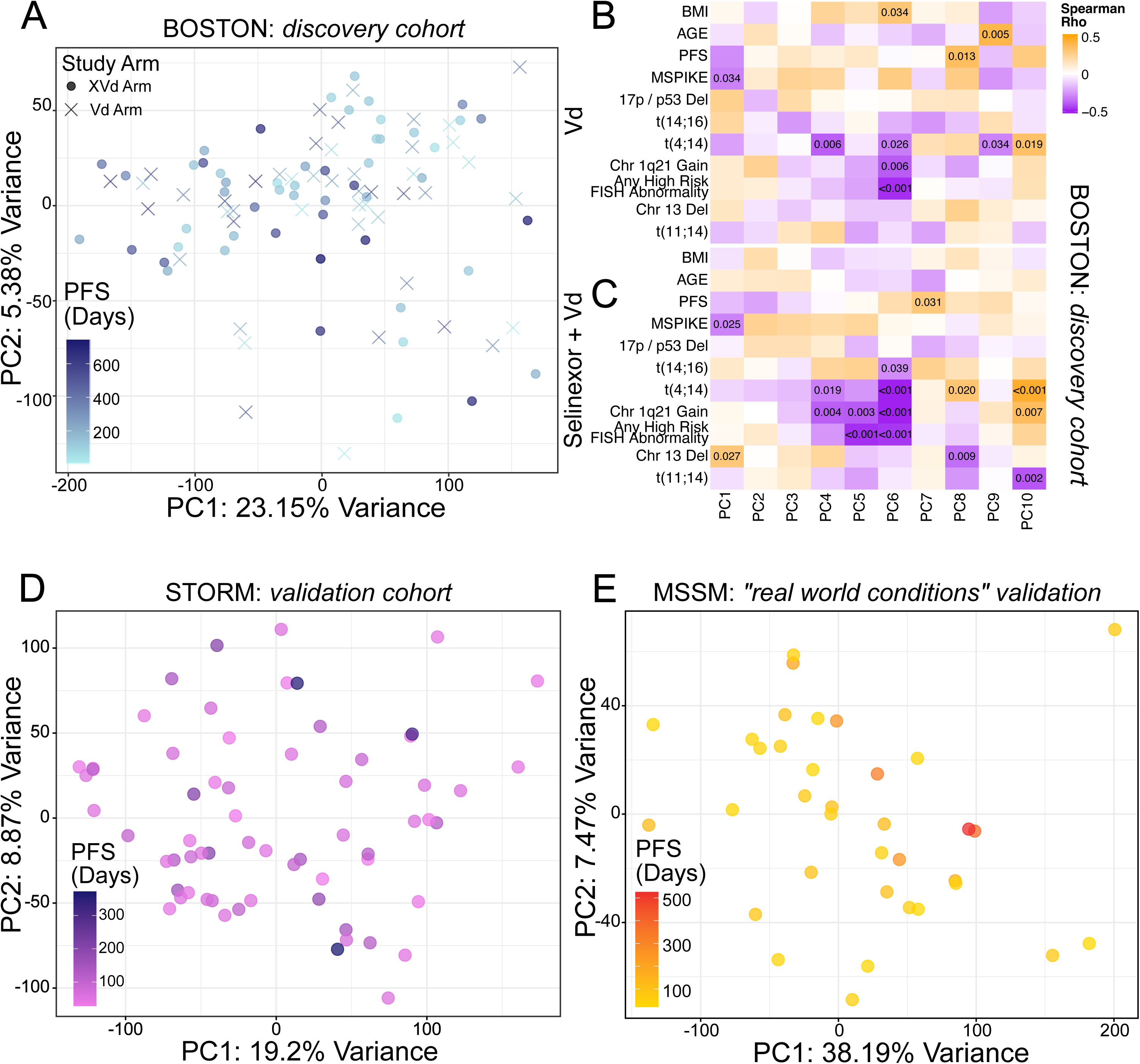
Normalized gene expression profiles of selinexor pre-treatment samples. A) PCA of BOSTON cohort normalized expression matrix colored by patient PFS in days. B) Spearman correlation of principal components with clinical variables, annotated with P values for correlations with P < 0.05, in BOSTON XVd and C) BOSTON Vd. D) PCA of STORM cohort normalized expression matrix, and E) MSSM cohort samples colored by patient PFS in days.

**Table S1.** Differential expression based signature definitions.

| Comparison Name | Responder Definition | XVd Arm:<br>N<br>Responders | XVd Arm:<br>N<br>Nonresponders | Vd Arm:<br>N<br>Responders | Vd Arm:<br>N<br>Nonresponders |
| --- | --- | --- | --- | --- | --- |
| PFS90 | PFS ≥ 90 Days | 43 | 10 | 34 | 13 |
| PFS120 | PFS ≥ 120 Days | 33 | 20 | 33 | 14 |
| PFS200 | PFS ≥ 200 Days | 23 | 30 | 25 | 22 |
| PFS300 | PFS ≥ 300 Days | 18 | 35 | 16 | 31 |
| PR | Best Response ≥ PR | 40 | 13 | 31 | 16 |
| VGPR | Best Response ≥ VGPR | 24 | 29 | 14 | 33 |
| PR_PFS120 | Best Response ≥ PR &<br>PFS ≥ 120 Days | 31 | 22 | 29 | 18 |
| VGPR_PFS120 | Best Response ≥ VGPR &<br>PFS ≥ 120 Days | 23 | 30 | 14 | 33 |
| VGPR_PFS300 | Best Response ≥ VGPR &<br>PFS ≥ 300 Days | 17 | 36 | 10 | 37 |

**Fig S2.**
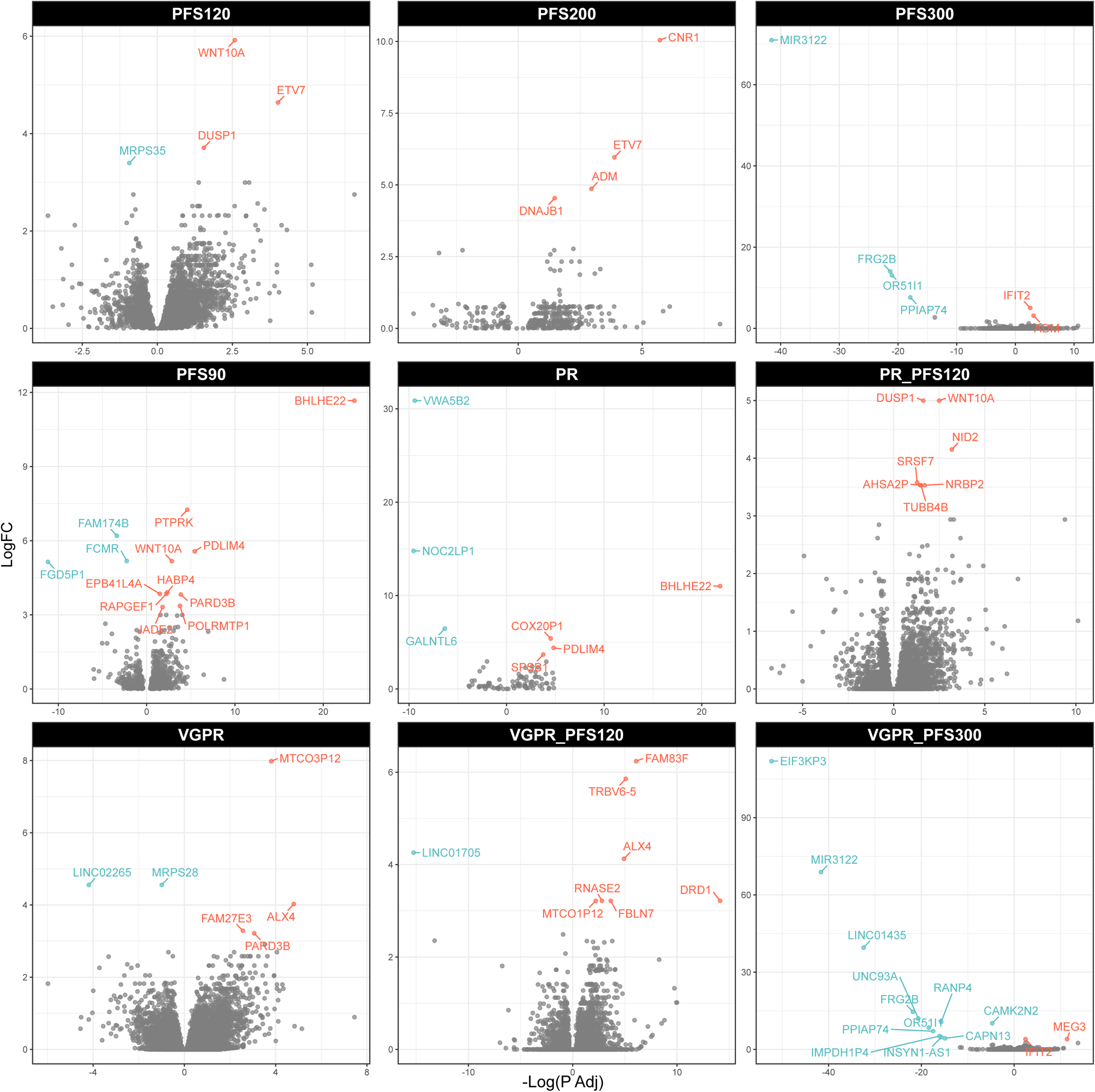
Differentially expressed genes in all unique BOSTON comparisons. Volcano plots showing all DE genes identified across response comparisons in the BOSTON XVd cohort. Gene symbols are labeled where Adj P < 0.05 in red for overexpression and blue for under-expression in responders to XVd therapy.

**Fig S3.**
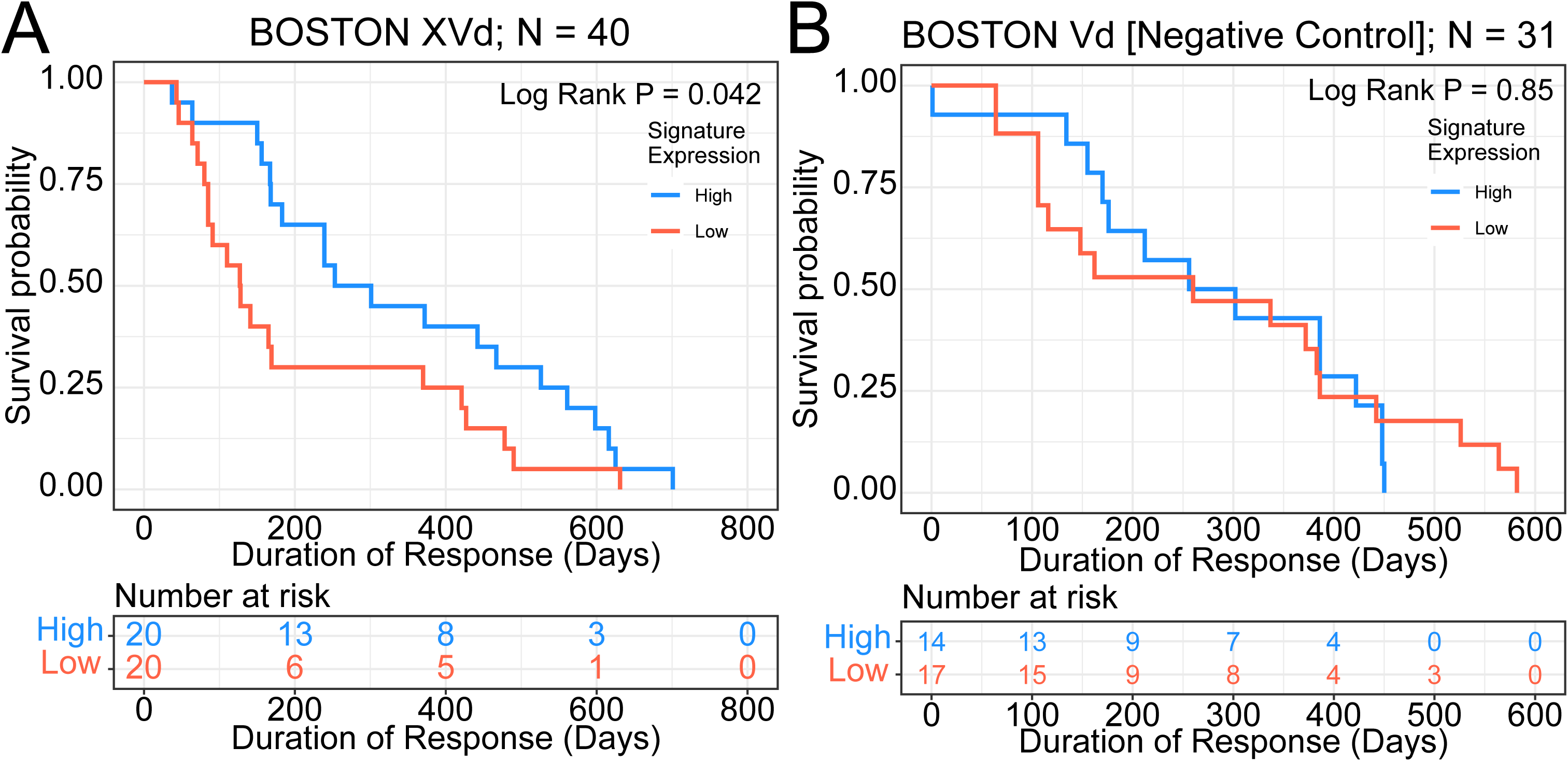
Signature expression tracks with longer duration of response in BOSTON XVd. A) Kaplan Meier curve of higher versus lower signature expression as a function of duration of response (DOR) in patients who achieved a partial response (PR) or better in A) the BOSTON XVd cohort shows a significant association, whereas there is no significant difference in B) patients in the BOSTON Vd cohort of non-selinexor based therapy.

**Fig S4.**
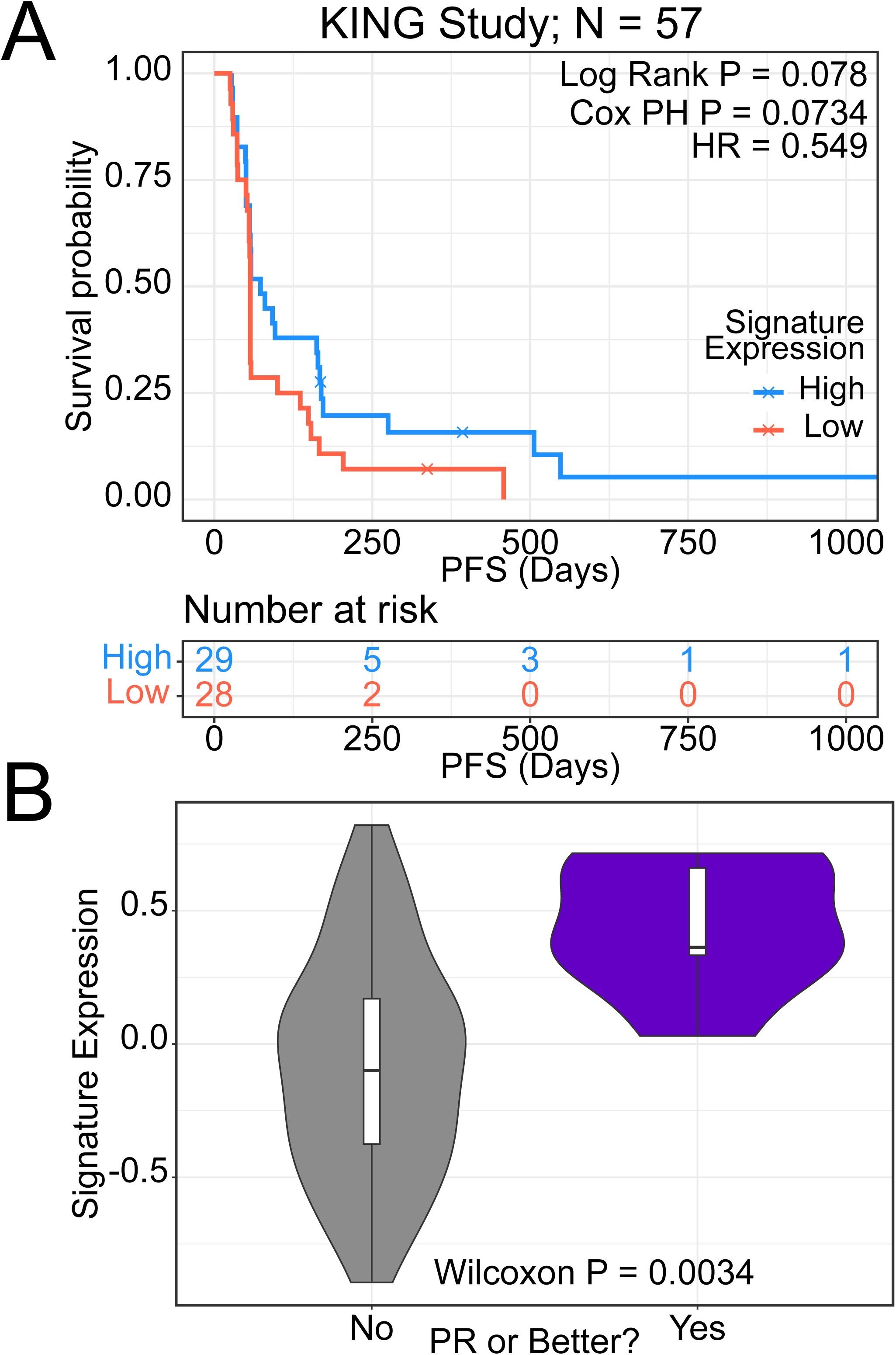
Signature expression predicts selinexor response in recurrent glioblastoma. A) Survival analysis of the KING recurrent glioblastoma cohort stratified by three-gene signature GSVA scores. B) Three gene signature GSVA scores in the KING cohort in responders (purple) versus non-responders (gray).

**Fig S5.**
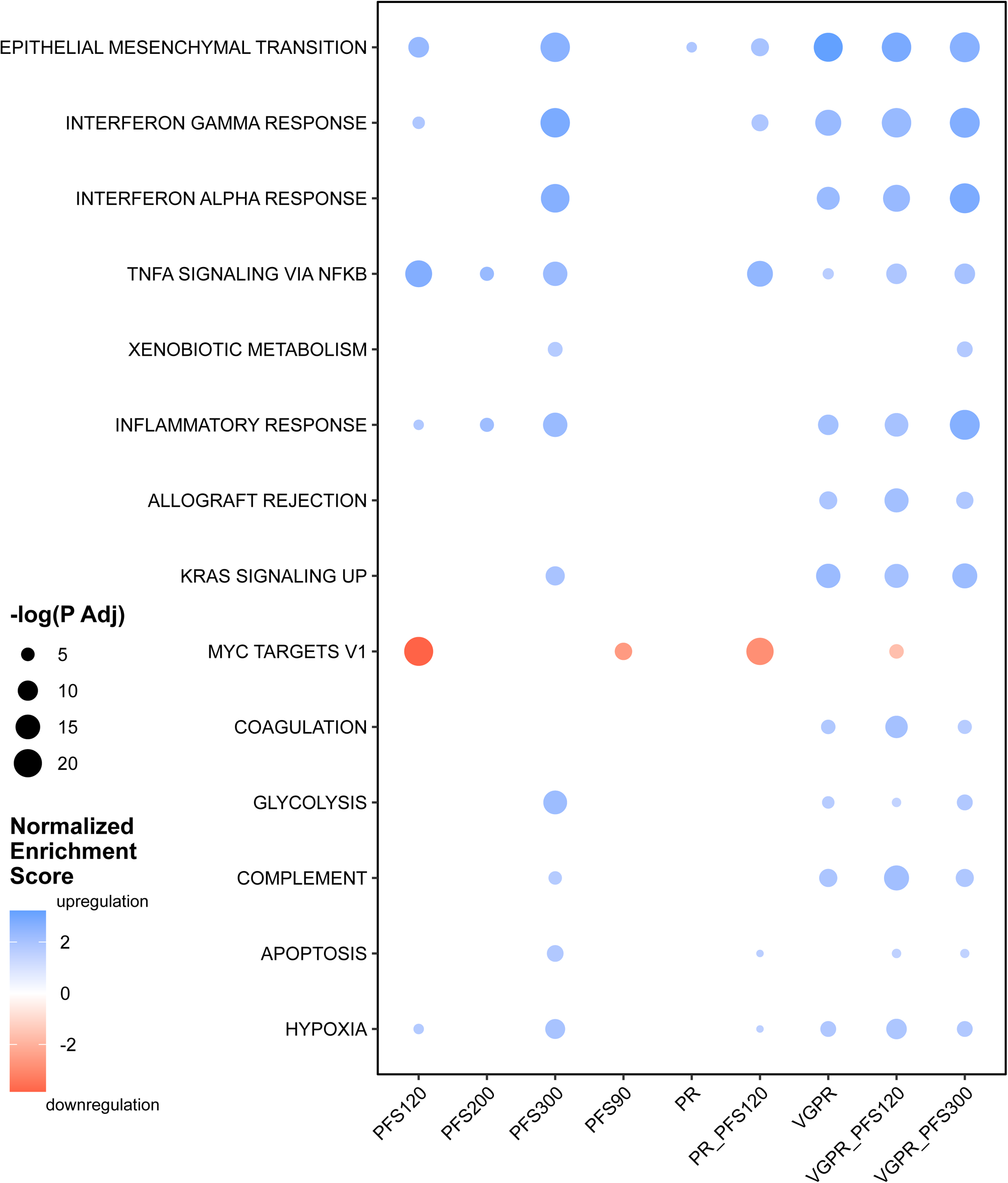
Functional enrichments in selinexor responders. GSEA enrichments in XVd responders across the various response cutoffs.

## REFERENCES

1. Grosicki S, Simonova M, Spicka I, et al: Once-per-week selinexor, bortezomib, and dexamethasone versus twice-per-week bortezomib and dexamethasone in patients with multiple myeloma (BOSTON): a randomised, open-label, phase 3 trial. The Lancet 396:1563–1573, 2020

2. Kalakonda N, Maerevoet M, Cavallo F, et al: Selinexor in patients with relapsed or refractory diffuse large B-cell lymphoma (SADAL): a single-arm, multinational, multicentre, open-label, phase 2 trial. The Lancet Haematology 7:e511–e522, 2020

3. Chari A, Vogl DT, Gavriatopoulou M, et al: Oral Selinexor–Dexamethasone for Triple-Class Refractory Multiple Myeloma. New England Journal of Medicine 381:727–738, 2019

4. Fung HYJ, Niesman A, Chook YM: An update to the CRM1 cargo/NES database NESdb. Mol Biol Cell 32:467–469, 2021

5. Chook YM, Fung HYJ: Atomic basis of CRM1-cargo recognition, release and inhibition. Semin Cancer Biol 0:52–61, 2014

6. Azmi AS, Uddin MH, Mohammad RM: The nuclear export protein XPO1 - from biology to targeted therapy. Nat Rev Clin Oncol, 2020

7. Tai Y-T, Landesman Y, Acharya C, et al: CRM1 inhibition induces tumor cell cytotoxicity and impairs osteoclastogenesis in multiple myeloma: molecular mechanisms and therapeutic implications. Leukemia 28:155–165, 2014

8. Neggers JE, Vanstreels E, Baloglu E, et al: Heterozygous mutation of cysteine528 in XPO1 is sufficient for resistance to selective inhibitors of nuclear export. Oncotarget 7:68842–68850, 2016

9. Bahlis NJ, Kotb R, Sebag M, et al: Selinexor in Combination with Bortezomib and Dexamethasone (SdB) Demonstrates Significant Activity in Patients with Refractory Multiple Myeloma (MM) Including Proteasome-Inhibitor Refractory Patients: Results of the Phase I Stomp Trial. Blood 128:977–977, 2016

10. Lagana A, Bhalla S, Aleman A, et al: A Machine Learning Approach Identifies a 30-Gene Model That Predicts Sensitivity to Selinexor in Multiple Myeloma. Blood 134:3101–3101, 2019

11. Kumar S, Paiva B, Anderson KC, et al: International Myeloma Working Group consensus criteria for response and minimal residual disease assessment in multiple myeloma. The Lancet Oncology 17:e328– e346, 2016

12. Lassman AB, Wen PY, van den Bent MJ, et al: A Phase II Study of the Efficacy and Safety of Oral Selinexor in Recurrent Glioblastoma. Clinical Cancer Research 28:452–460, 2022

13. Liberzon A, Subramanian A, Pinchback R, et al: Molecular signatures database (MSigDB) 3.0. Bioinformatics 27:1739–1740, 2011

14. Laganà A, Beno I, Melnekoff D, et al: Precision Medicine for Relapsed Multiple Myeloma on the Basis of an Integrative Multiomics Approach. JCO Precision Oncology 1–17, 2018

15. Dobin A, Davis CA, Schlesinger F, et al: STAR: ultrafast universal RNA-seq aligner. Bioinformatics 29:15–21, 2013

16. Liao Y, Smyth GK, Shi W: featureCounts: an efficient general purpose program for assigning sequence reads to genomic features. Bioinformatics 30:923–930, 2014

17. Ritchie ME, Phipson B, Wu D, et al: limma powers differential expression analyses for RNA-sequencing and microarray studies. Nucleic Acids Research 43:e47, 2015

18. Hoffman GE, Schadt EE: variancePartition: interpreting drivers of variation in complex gene expression studies. BMC Bioinformatics 17:483, 2016

19. Leek JT, Johnson WE, Parker HS, et al: The sva package for removing batch effects and other unwanted variation in high-throughput experiments. Bioinformatics 28:882–883, 2012

20. Love MI, Huber W, Anders S: Moderated estimation of fold change and dispersion for RNA-seq data with DESeq2. Genome Biology 15:550, 2014

21. Korotkevich G, Sukhov V, Budin N, et al: Fast gene set enrichment analysis [Internet]060012, 2021[cited 2022 Feb 17] Available from: https://www.biorxiv.org/content/10.1101/060012v3

22. Hänzelmann S, Castelo R, Guinney J: GSVA: gene set variation analysis for microarray and RNA-Seq data. BMC Bioinformatics 14:7, 2013

23. Franks JM, Cai G, Whitfield ML: Feature specific quantile normalization enables cross-platform classification of molecular subtypes using gene expression data. Bioinformatics 34:1868–1874, 2018

24. Miloudi H, Bohers É, Guillonneau F, et al: XPO1E571K Mutation Modifies Exportin 1 Localisation and Interactome in B-Cell Lymphoma. Cancers (Basel) 12:2829, 2020

25. Baumhardt JM, Walker JS, Lee Y, et al: Recognition of nuclear export signals by CRM1 carrying the oncogenic E571K mutation. Mol Biol Cell 31:1879–1891, 2020

26. Leaderer D, Hoffman AE, Zheng T, et al: Genetic and epigenetic association studies suggest a role of microRNA biogenesis gene exportin-5 (XPO5) in breast tumorigenesis. Int J Mol Epidemiol Genet 2:9– 18, 2011

27. Martini S, Zuco V, Tortoreto M, et al: miR-34a-Mediated Survivin Inhibition Improves the Antitumor Activity of Selinexor in Triple-Negative Breast Cancer. Pharmaceuticals (Basel) 14:523, 2021

28. Chen Q, Gong B, Mahmoud-Ahmed AS, et al: Apo2L/TRAIL and Bcl-2–related proteins regulate type I interferon–induced apoptosis in multiple myeloma. Blood 98:2183–2192, 2001

29. Agnarelli A, Chevassut T, Mancini EJ: IRF4 in multiple myeloma—Biology, disease and therapeutic target. Leukemia Research 72:52–58, 2018

30. Dufva O, Pölönen P, Brück O, et al: Immunogenomic Landscape of Hematological Malignancies. Cancer Cell 38:380-399.e13, 2020

31. Liao Y, Ke X, Deng T, et al: The Second-Generation XPO1 Inhibitor Eltanexor Inhibits Human Cytomegalovirus (HCMV) Replication and Promotes Type I Interferon Response. Front Microbiol 12:675112, 2021

32. Pezzè L, Meškytė EM, Forcato M, et al: ETV7 regulates breast cancer stem-like cell features by repressing IFN-response genes. Cell Death Dis 12:1–14, 2021

