## Supplementary figures and images for "A three gene signature predicts response to selinexor in multiple myeloma"

### Supplemental Figure 1

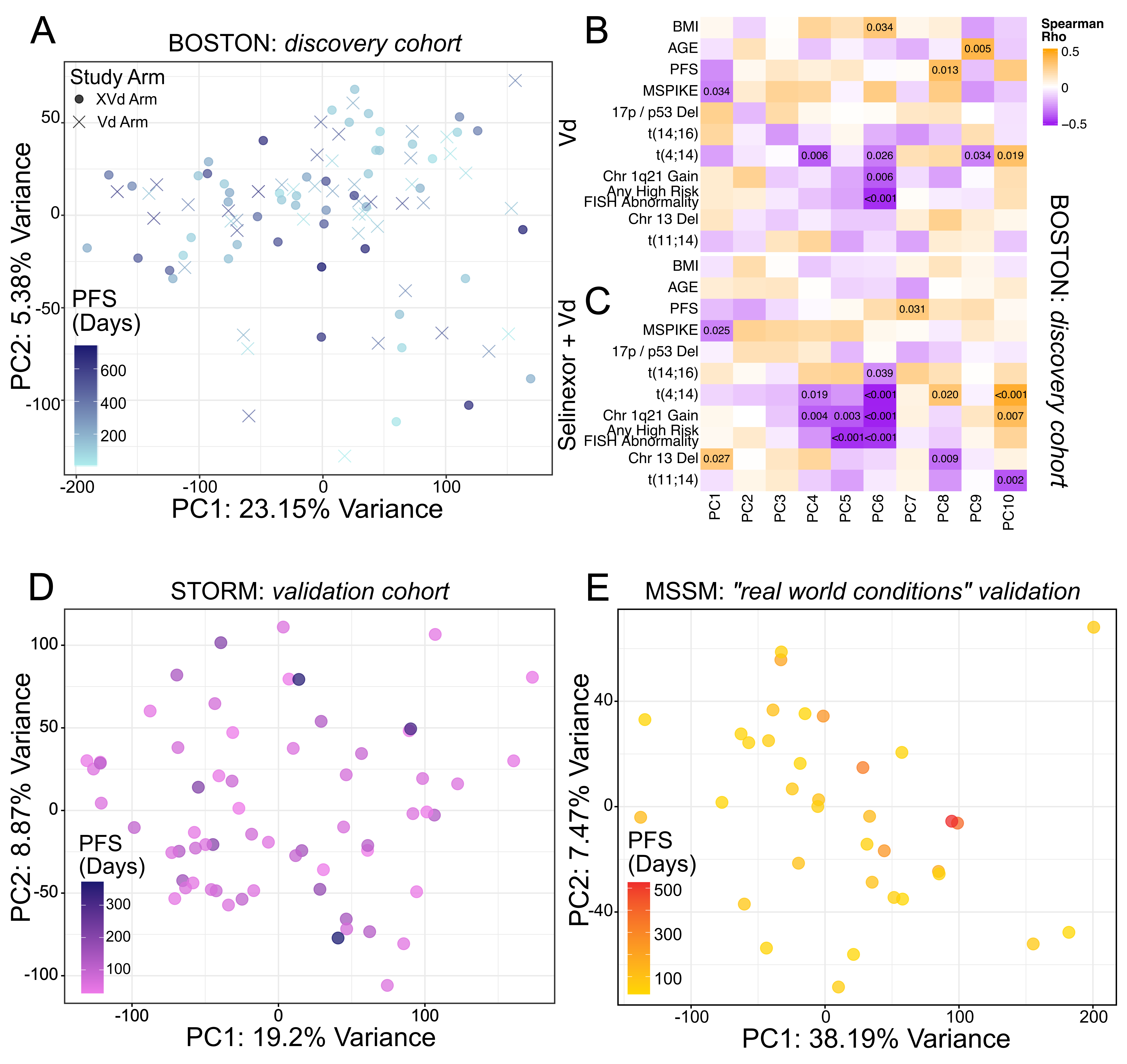

### Supplemental Figure 2

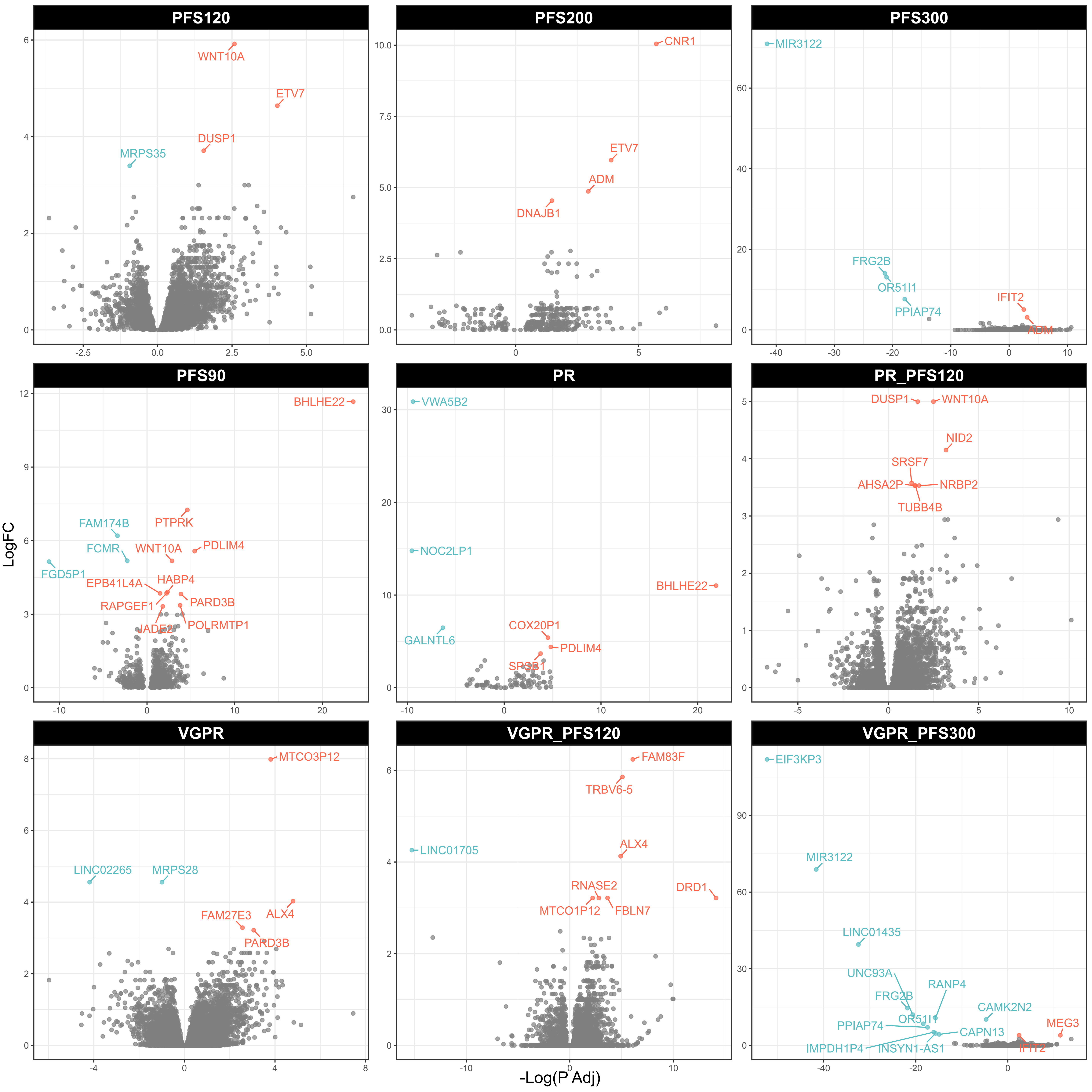

### Supplemental Figure 3

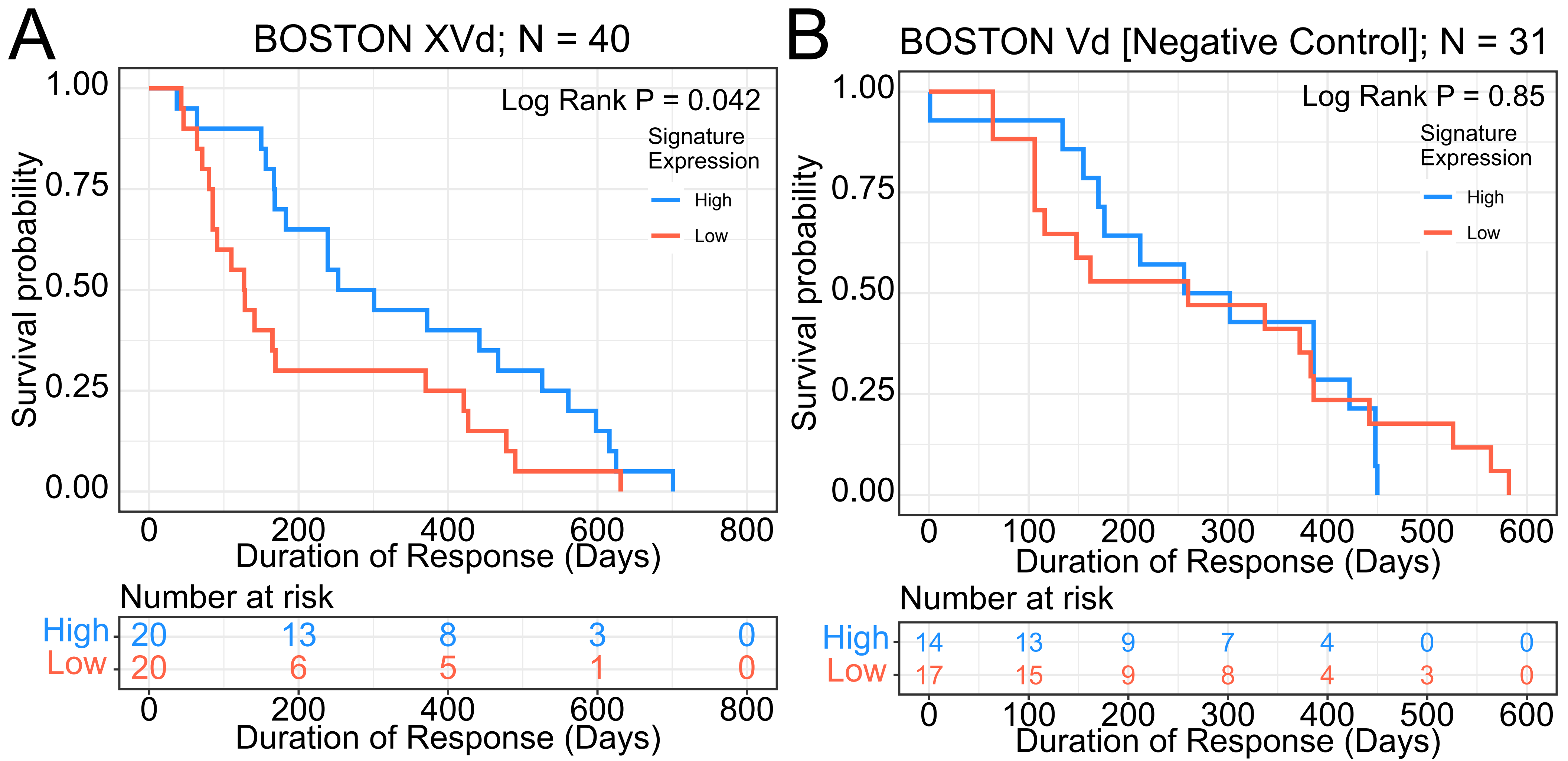

### Supplemental Figure 4

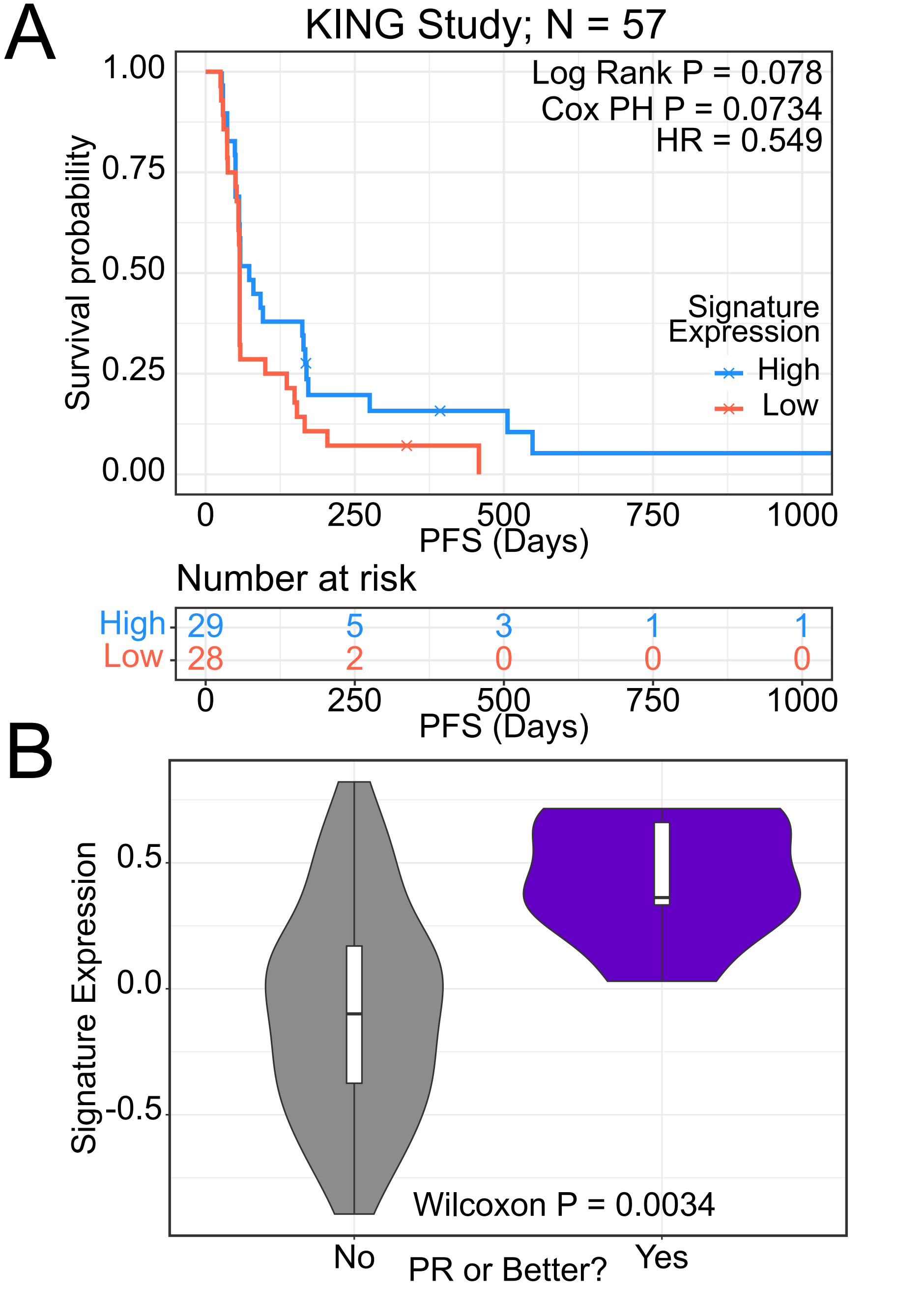

### Supplemental Figure 5

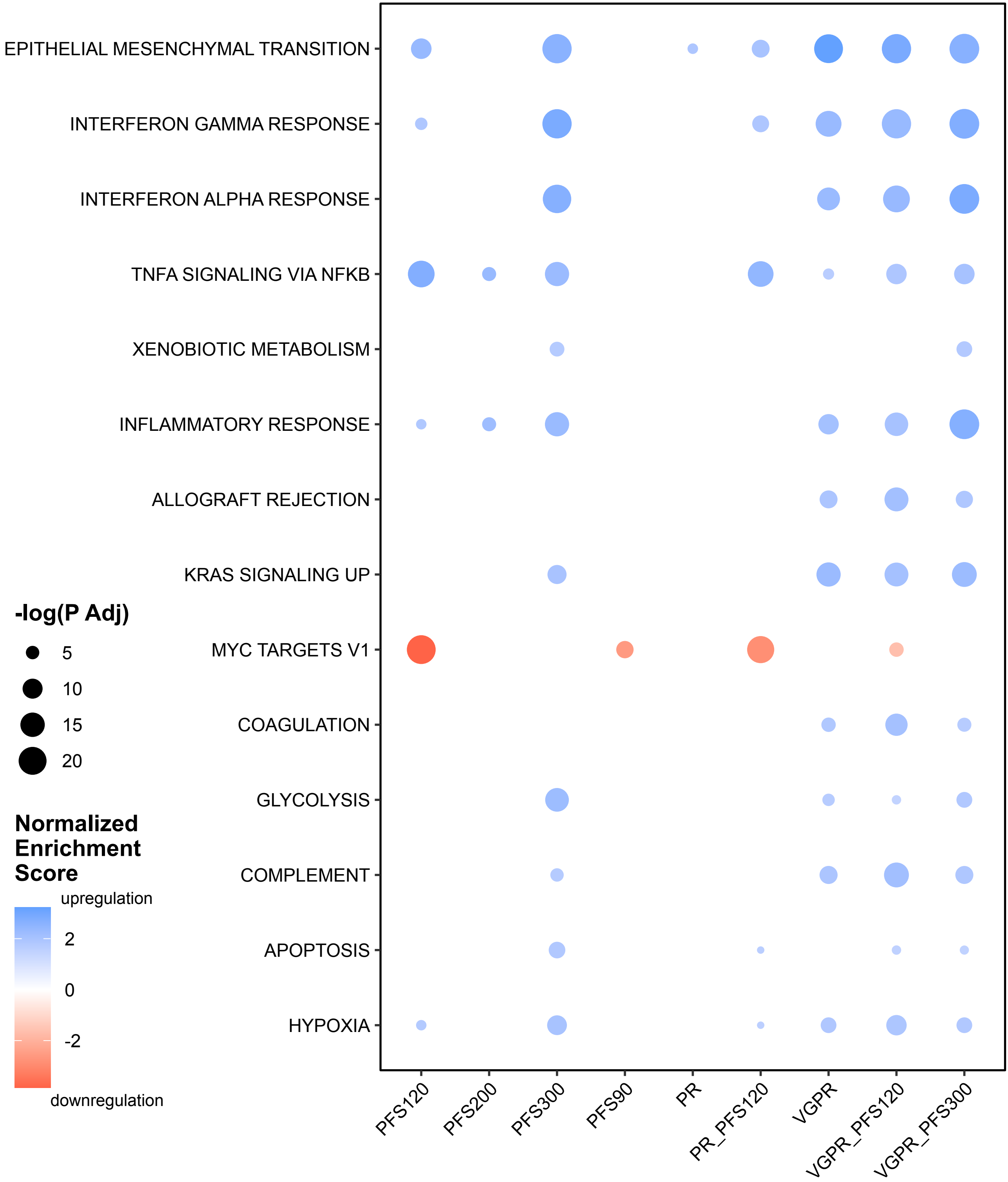
